# Diabetes is associated with increased nocturnal respiratory rate

**DOI:** 10.64898/2026.06.16.26355548

**Authors:** Keshav S. Gupta, Raimon Padrós-Valls, Nicholas Harrington, David Torres Barba, Kevin R. King

## Abstract

**Background and Objective:** Diabetes mellitus (DM) causes autonomic neuropathy, which may alter nocturnal respiratory rate (NRR). To test the association between DM and NRR, we analyzed elective polysomnograms of four large observational cohorts.

**Research Design and Methods:** We performed cross-sectional analysis of over 25,000 individuals with polysomnograms (PSGs) from the Sleep Heart Health Study (SHHS), Hispanic Community Health Study/Study of Latinos (HCHS/SOL), Osteoporotic Fractures in Men Study (MrOS), and Wisconsin Sleep Cohort (WSC). Patient-level NRRs were derived from inductance plethysmography waveforms. DM status was determined by self-report, physician diagnosis, medication use, or laboratory values, depending on the cohort. We related DM and NRR (continuous and dichotomized) using logistic regression models and adjusted for potential confounders. Cohort-specific results were combined using random-effects meta-analysis.

**Results:** Meta-analysis of unadjusted models showed a pooled odds ratio (OR) of 1.10 (95% CI:1.04–1.17) for each breath-per-minute (brpm) increase in NRR. This association remained significant after multivariable adjustment (OR:1.06, 95% CI:1.02–1.11). Dichotomized analyses similarly showed higher odds of DM across dichotomization thresholds ranging from 15 to 21 brpm. At a threshold of 18 brpm, the unadjusted pooled OR was 1.77 (95% CI:1.23–2.55, P=0.0022), and the adjusted OR was 1.49 (95% CI:1.10–2.02, P=0.0098).

**Conclusions:** Clinically stable outpatients with elevated NRR have an increased prevalence of DM. Additional studies are needed to investigate whether the mechanism is autonomic neuropathy and whether monitoring NRR can detect early complications of DM.

**ARTICLE HIGHLIGHTS:**

- **Why did we undertake this study?** Diabetes mellitus (DM) is linked to autonomic neuropathy, and nocturnal respiratory rate (NRR) is regulated by the autonomic nervous system; yet the relationship between NRR and DM is unknown.
- **What specific question did we want to answer?** Is NRR associated with DM? To answer this, we derived NRR and DM status from 25,000+ PSGs.
- **What did we find?** Individuals with elevated NRR have higher DM prevalence even after adjusting for confounding variables. Additionally, groups with DM had a significantly higher median NRR compared to those without DM.
- **What are the implications of our findings?** If the mechanism of association between NRR and DM involves autonomic neuropathy, then NRR may represent an easy-to-longitudinally-monitor biomarker for development of DM complications.

## INTRODUCTION

Diabetes Mellitus (DM) is a systemic disease that causes neurovascular complications that affect nearly every organ^1-4^. Biomarkers of acute and subacute glycemic control such as blood glucose and glycated hemoglobin are well established and routinely monitored. In contrast, biomarkers of DM-associated complications are monitored infrequently, inconsistently, or not at all^5^. Autonomic neuropathy is a complication of diabetes that represents an attractive target for continuous monitoring because it effects cardiopulmonary physiology, which is increasingly measurable by wearables and non-contact ambient sensors^6-8^. While substantial effort has been devoted to studying the relationship between diabetes and cardiac autonomic dysfunction via heart rate variability, little is known about how diabetes affects respiratory biomarkers, which are also modulated by the autonomic nervous system. Studies relating DM to respiratory rate typically focus on acutely decompensated hospitalized patients with diabetic ketoacidosis in whom rapid shallow breathing serves as respiratory compensation for primary metabolic acidosis. However, little is known about the relationship between respiratory rate in clinically stable outpatients with diabetes. Here, we examine the relationship between DM and respiratory rate in ambulatory patient undergoing elective outpatient sleep studies.

Respiratory rate is considered a fundamental vital sign, yet it is most often measured imprecisely by manually counting the number of breaths a person takes in 15 to 30 seconds while awake in clinic. As a result, it is largely ignored except at its extremes. In awake individuals, respiratory rate is highly variable and difficult to interpret because it is dominated by conscious inputs such as effort, physical activity, and emotional stress^9^. However, during sleep, conscious confounders subside, allowing the nocturnal respiratory rate (NRR) to reflect fundamental underlying physiology that may serve as a high-quality biomarker of the central pattern generator and autonomic nervous system tone^10, 11^. We therefore derived patient-level longitudinal NRRs from polysomnogram waveforms of 4 large observational cohorts, and related them to the presence or absence of DM.

## RESEARCH DESIGN AND METHODS

### Study Design

The association between DM and NRR was cross-sectionally examined in four large, publicly available datasets. For individuals with multiple PSGs, just the first one was analyzed. Within each dataset, univariate and multivariate logistic regression models were used to assess the odds ratio (OR) and 95% confidence interval (CI) for DM in relation to patient level median NRR, treated either as a continuous variable or dichotomized. The results from the individual datasets were combined using a random-effects meta-analytic approach (DerSimonian-Laird) to obtain pooled odds ratios (ORs) and confidence intervals (CIs).

### Study Population

We analyzed data from the following four observational cohorts (Figure 1, Table 1). The **Sleep Heart Health Study (SHHS)** enrolled 6,441 men and women aged ≥40 years. We excluded 1,281 individuals due to poor-quality PSG or missing covariates resulting in a final analytic sample of 5,160 participants. DM status was based on self-reporting during health interviews (detailed in the study’s protocol).^12-14^ The **Hispanic Community Health Study / Study of Latinos (HCHS/SOL)** enrolled 16,415 adults (18–74 years) who self-identified as Hispanic/Latino, through a stratified multistage area probability sample in four communities. We excluded 5,270 individuals due to poor-quality PSG or missing covariates, leaving a final analytic sample of 11,145 participants. DM status was determined from questionnaires, medication inventories, and blood tests, and categorized participants as “normal,” “Abnormal Fasting Glucose,” or “Diabetes.” For the current analysis, “normal” and “Abnormal Fasting Glucose” were combined into a single non-DM category.^14-17^ The **Osteoporotic Fractures in Men Study (MrOS)** consists of 5,994 community-dwelling men aged 65 years or older, of which 3,135 participants were enrolled in a polysomnogram sub-cohort. We excluded 617 individuals due to poor-quality PSG or missing covariates leaving 2,518 participants for analysis. DM status was determined from medical history, fasting glucose, and medication use. As with HCHS/SOL, “normal” and “Abnormal Fasting Glucose” were combined into one non-DM group.^14, 16, 18-20^ The **Wisconsin Sleep Cohort (WSC)** enrolled 1,522 Wisconsin state employees and yielded 1,123 participants with elective overnight polysomnograms and associated electronic medical record data.^14, 16, 21^ We excluded 120 individuals for missing covariates, leaving 1,003 participants for analysis. DM status was determined by self-reported physician diagnosis.

**Figure 1:**
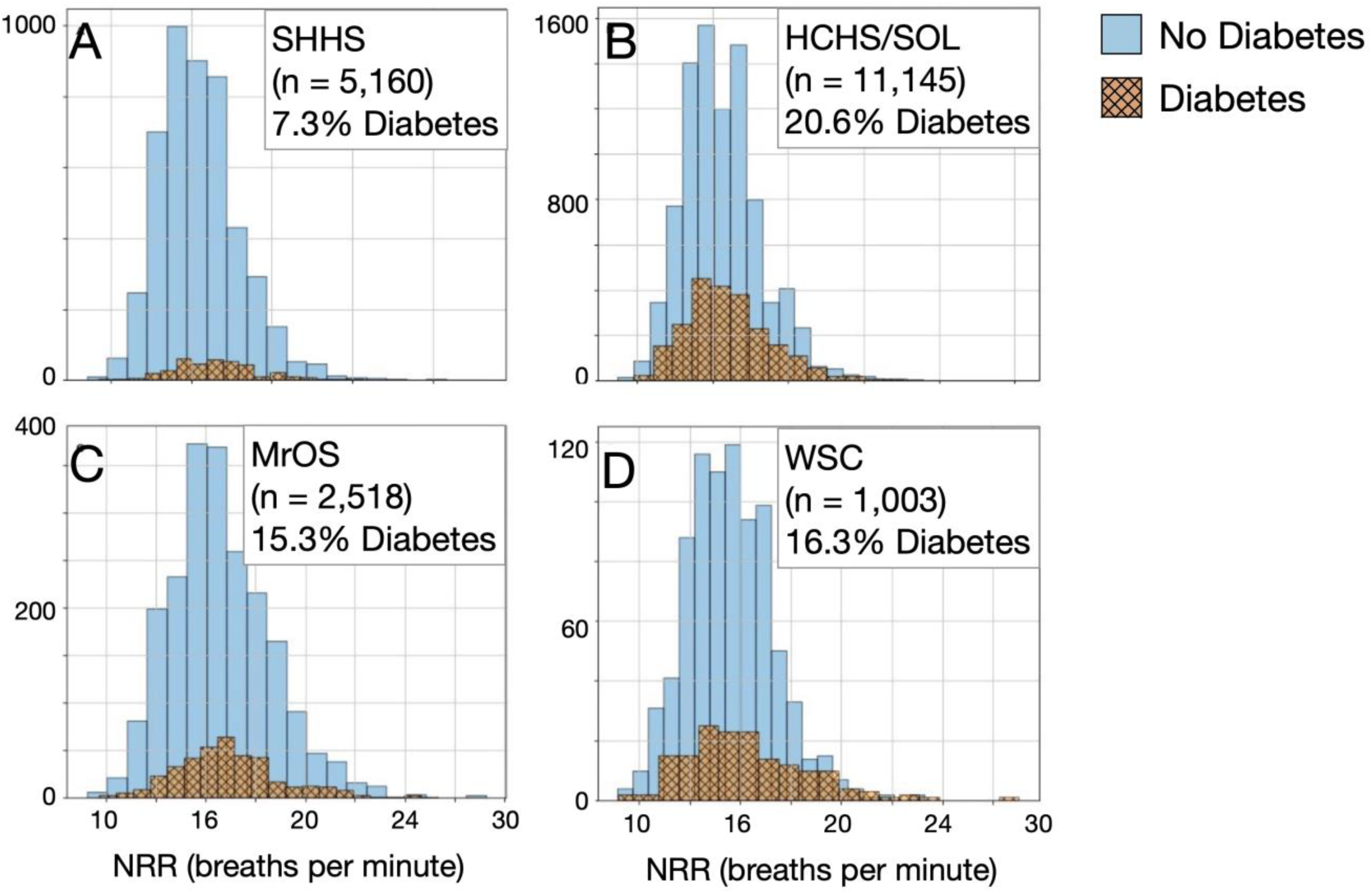
Histograms of NRR for segmented based on diabetes status. **A) SHHS:** Total participants = 5,160; Diabetics = 376 (7.3%), Non-diabetics = 4,784 (92.7%). NRR of Non-diabetics: mean = 15.5, median = 15.4; NRR of Diabetics: mean = 16.6, median = 16.2. **B) HCHS/SOL:** Total participants = 11,145; Diabetics = 2,293 (20.6%), Non-diabetics = 8,852 (79.4%). NRR of Non-diabetics: mean = 15.6, median = 15.4; NRR of Diabetics: mean = 16.0, median = 15.6. **C) MrOS:** Total participants = 2,518; Diabetics = 384 (15.3%), Non-diabetics = 2,134 (84.7%). NRR of Non-diabetics: mean = 15.7, median = 15.5; NRR of Diabetics: mean = 16.1, median = 15.9. **D) WSC:** Total participants = 1,003; Diabetics = 163 (16.3%), Non-diabetics = 840 (83.7%). NRR of Non-diabetics: mean = 14.5, median = 14.4; NRR of Diabetics: mean = 15.3, median = 14.8.

**Table 1:** Demographics, respiratory covariates, and NRR for DM and non-DM subsets of the 4 observational data sets.

|  |  | SHHS |  |  | HCHS |  |  | MROS |  |  | WSC |  |  |
| --- | --- | --- | --- | --- | --- | --- | --- | --- | --- | --- | --- | --- | --- |
|  |  | DM | Non-DM | p-value | DM | Non-DM | p-value | DM | Non-DM | p-value | DM | Non-DM | p-value |
| **NRR (brpm)** |  | 16.2 (14.8-18.2) | 15.4 (13.6-17.1) | 1.14E-14 | 15.6 (14.0-17.7) | 15.4 (13.8-17.1) | 2.75E-06 | 15.9 (14.3-17.5) | 15.48 (13.91-17.14) | 6.52E-03 | 14.9 (13.3-17.1) | 14.4 (13.0-15.9) | 6.04E-03 |
| **age (years)** |  | 70 (63-76) | 63 (55-72) | 2.57E-25 | 56 (49-62) | 46 (34-54) | 2.03E-266 | 75 (72-79) | 76 (72-80) | 1.31E-01 | 65 (60-71) | 62 (55-68) | 8.75E-06 |
| **bmi** |  | 29.3 (26.5-33.4) | 27.4 (24.6-30.6) | 6.29E-15 | 30.9 (27.6-35.3) | 28.4 (25.3-32.1) | 3.71E-91 | 28 (26-32) | 26 (24-29) | 1.31E-01 | 33.7 (29.8-40.7) | 29.6 (26.3-34.3) | 1.38E-12 |
| **Sex (Female)** |  | 44.9 | 53.2 | 2.56E-03 | 60.7 | 59.7 | 4.17E-01 | 0 | 0 | 5.61E-18 | 50.9 | 45.8 | 2.69E-01 |
| **Race** | White | 80.3 | 86.6 | 2.89E-07 | - | - | - | 88 | 91.7 | 2.46E-03 | 95.7 | 97.7 | - |
|  | African-American | 14.6 | 6.9 |  |  |  |  | 5.7 | 2.7 |  | 1.8 | 1.2 |  |
|  | Asian | - | - |  |  |  |  | 3.1 | 2.7 |  | 0.6 | 0.6 |  |
|  | Hispanic | 84.9 | 89.1 |  | 100 | 100 | - | 1.1 | 2.1 | - | 1.8 | 0.5 | - |
|  | Other | 5.1 | 6.4 |  |  |  |  | 2.1 | 0.8 |  |  |  |  |
| **Smoking** | Never | 45.2 | 47.7 | 5.57E-02 | 57.5 | 62.3 | 1.24E-19 | 37 | 40.5 | 2.19E-01 | 41.1 | 51.3 | 6.47E-04 |
|  | Past | 6.6 | 9.5 |  | 27.1 | 18.5 | - | 61.7 | 57.4 |  | 54.6 | 39.3 | - |
|  | Current | 48.1 | 42.8 |  | 15.4 | 19.2 | - | 1.3 | 2.1 |  | 4.3 | 9.4 | 2.46E-01 |
| **Sleep Apnea** | None (AHI < 5) | 9 | 18.4 | 1.34E-08 | 50.9 | 73.9 | 2.44E-107 | 7.3 | 10.4 | 2.60E-02 | 39.3 | 45.5 | 1.25E-02 |
|  | Mild (5< AHI < 15) | 34 | 39.3 |  | 27.9 | 17.2 | - | 29.9 | 34.2 |  | 25.2 | 30 |  |
|  | Moderate (15< AHI < 30) | 33.5 | 27 |  | 12.6 | 5.9 | - | 34.1 | 32 |  | 19 | 15.5 | - |
|  | Severe (AHI >30) | 23.4 | 15.3 |  | 8.6 | 3.1 | - | 28.6 | 23.3 |  | 16.6 | 9 | - |

### Nocturnal Respiratory Rate Derivation

NRR was derived from the chest belt impedance plethysmography signal recorded during the overnight PSG. To reduce the effect of sleep onset and waking, the first and last hours of the night were discarded. Inspiratory onsets were identified by performing peak detection, and the respiratory rate was computed every minute with a 5-minute sliding window. Windows were excluded if the respiratory rate exceeded 40 brpm or fell below 6 brpm or if any breath-to-breath interval within the window exceeded 50 brpm or fell below 5 brpm. The subject’s median respiratory rate was calculated across all remaining windows.

### Statistical Analysis

All computations were performed in Python (version 3.12.2) using the pandas (version 2.2.2), numpy (version 1.26.4), statsmodels (version 0.14.2), and scipy (version 1.11.4) libraries.

### Covariates

Logistic regression models were adjusted for covariates that could potentially confound the relationship between NRR and DM: age (years), sex (Male/Female), body mass index (BMI, kg/m²), smoking history (never/past/current), sleep apnea severity (none/mild/moderate/severe), and race/ethnicity (White, African-American, Hispanic, Asian, Other). Age and BMI were modeled as continuous variables. Sex was included as a binary variable (Male/Female). Sleep apnea severity was categorized based on the apnea–hypopnea index (AHI): none (AHI < 5), mild (5 ≤ AHI < 15), moderate (15 ≤ AHI < 30), and severe ( AHI ≥ 30); this variable, along with smoking history and race/ethnicity, was included in the model using one-hot encoding to accommodate its multi-categorical structure. All covariates were used as recorded in the source datasets. In cohort-specific models, covariate inclusion was adapted based on the demographic composition of each cohort. For example, the MrOS dataset included only male participants; therefore, sex was not included as a covariate in models for this cohort. Similarly, the HCHS/SOL cohort consisted exclusively of Hispanic participants, and as such, race/ethnicity was not included in HCHS/SOL-specific models. Additionally, race/ethnicity category availability varied across cohorts: SHHS did not include the ‘Asian’ category, and WSC did not include the ‘Other’ category. Descriptive statistics for covariate distributions in DM and non-DM participants are presented in Table 1. Further details about variable definitions and data collection protocols in each cohort can be found at https://sleepdata.org. Variance Inflation Factor (VIF) analyses were performed to check for multicollinearity. A cutoff of 5 was used to reduce the risk of overestimated confidence intervals or Type II errors.

### Descriptive Statistics

Within each dataset, participants were categorized into DM and non-DM groups. Categorical variables were expressed as percentages, while continuous variables were summarized as median (interquartile range) (**Table 1**). To visually compare the distribution of NRR between DM and non-DM individuals, both groups were plotted together (**Figure 1**). Group differences in covariates were assessed using chi-square tests for categorical variables. For continuous variables, the Mann-Whitney U test was applied to test for differences (**Table 1**).

### NRR as a Continuous Predictor

First, the individual’s median NRR was treated as a continuous variable. Univariate logistic regression produced unadjusted ORs and 95% CIs for DM for a 1 breath per minute (brpm) increase in NRR. Multivariable logistic regression models were then employed, adjusting for the covariates, to produce adjusted ORs and 95% CIs. This procedure was repeated for each of the four datasets.

### NRR as a Binary Predictor

Because no established clinical cutoff exists for dichotomizing NRR, we dichotomized the median NRR variable at seven thresholds (15- 21 brpm). For each cutoff, the sample was split into NRR ≥ cutoff vs. NRR < cutoff, and both unadjusted and adjusted logistic regression models were run in each dataset, generating ORs and 95% CIs for DM. The cutoff of 15 was chosen because it approximated the me an NRR across the datasets, and 21 was included as it corresponded to about the 99th percentile. Since continuous variables are often dichotomized in clinical medicine for practicality, we examined the association at multiple different cut-offs, however, we only reported OR and 95% CI for the 18 brpm cut-off.

### Meta-Analysis

For both continuous and binary analyses, each dataset produced unadjusted and adjusted ORs with corresponding 95% CIs. These were then combined via a random-effects meta-analysis using the DerSimonian and Laird method. The random effects model was chosen since the *I*^2^ Statistic was >50%, indicating a significant heterogeneity between datasets, as was expected. For the continuous NRR measure, each dataset yields a single unadjusted and adjusted OR and 95% CIs. These were combined using the DerSimonian and Laird method to provide pooled OR and CIs. For the dichotomization study, each dataset yields 7 adjusted and 7 unadjusted ORs and CIs corresponding to the 7 chosen cutoff points (15-21 brpm). These are combined via the DerSimonian and Laird method to provide pooled adjusted and unadjusted ORs and CIs at the 7 cut-off points.

### Data and Resource Availability

The data used in this study is available from the National Sleep Research Resource (NSRR) upon request.

## RESULTS

### NRR Distributions and Group Comparisons

To investigate the relationship between DM and NRR, we analyzed 27,114 PSGs from 4 large observational cohorts with polysomnography substudies (6,441 SHHS; 16,415 HCHS; 3,135 MrOS; 1,123 WSC). We excluded 7,288 (26.9%) of participants based on poor quality PSG waveforms or missing covariate metadata, leaving 19,826 PSGs for final analysis (5,160 SHHS; 11,145 HCHS; 2,518 MrOS; 1,003 WSC). **Table 1** details the diversity of demographics and covariates segmented by DM and non-DM subgroups for each cohort. At one extreme, MROS subjects were all male, were older (mid-70s), predominantly white, and had the lowest BMIs; at the other, HCHS were the 60% female, approximately 2 to 4 decades younger, and 100% Hispanic. **Figure 1A-D** shows the distribution of NRR and burden of DM in participants across each dataset. The proportion of DM across the datasets ranged from 7.3% in SHHS to 20.6% in HCHS. In all 4 cohorts, the mean and median NRR were higher in the DM group than in the non-DM group. Across all datasets, the DM group had a statistically significant higher NRR (p < 0.05) under the nonparametric Mann-Whitney U tests.

### Continuous NRR

Univariate and multivariate logistic regression models were used to evaluate continuous NRR as a predictor of prevalent diabetes mellitus (DM) in each cohort. In the unadjusted models, NRR was significantly associated with increased odds of DM across all four cohorts: SHHS (OR = 1.15, 95% CI: 1.11–1.20, *P* = 2.7 × 10⁻¹⁴), HCHS/SOL (OR = 1.05, 95% CI: 1.03–1.07, *P* = 4.8 × 10⁻⁹), MrOS (OR = 1.06, 95% CI: 1.02–1.11, *P* = 0.0047), and WSC (OR = 1.15, 95% CI: 1.08–1.23, *P* = 4.1 × 10⁻⁵). In the adjusted models, controlling for age, sex, BMI, smoking history, sleep apnea severity, and race when applicable, the association remained statistically significant in SHHS (OR = 1.11, 95% CI: 1.06 –1.15, *P* = 3.5 × 10⁻⁷), HCHS/SOL (OR = 1.04, 95% CI: 1.03–1.06, *P* = 4.4 × 10⁻⁶), and WSC (OR = 1.12, 95% CI: 1.04–1.21, *P* = 0.0021). In MrOS, the association was attenuated and no longer statistically significant (OR = 1.03, 95% CI: 0.99–1.08, *P* = 0.13), potentially reflecting limited statistical power or the lack of sex diversity in this all-male cohort. **Figure 2A** shows the random-effects meta-analysis of univariate logistic regression results, revealing a pooled OR of 1.10 (95% CI: 1.04–1.16, P = 3.5 × 10⁻⁴). **Figure 2B** shows the multivariate meta-analysis, indicating a slightly attenuated pooled OR of 1.07 (95% CI: 1.03–1.11, P = 3.0 × 10⁻⁴). These findings suggest that a higher continuous NRR is significantly associated with prevalent DM, even after adjusting for relevant covariates.

**Figure 2.**
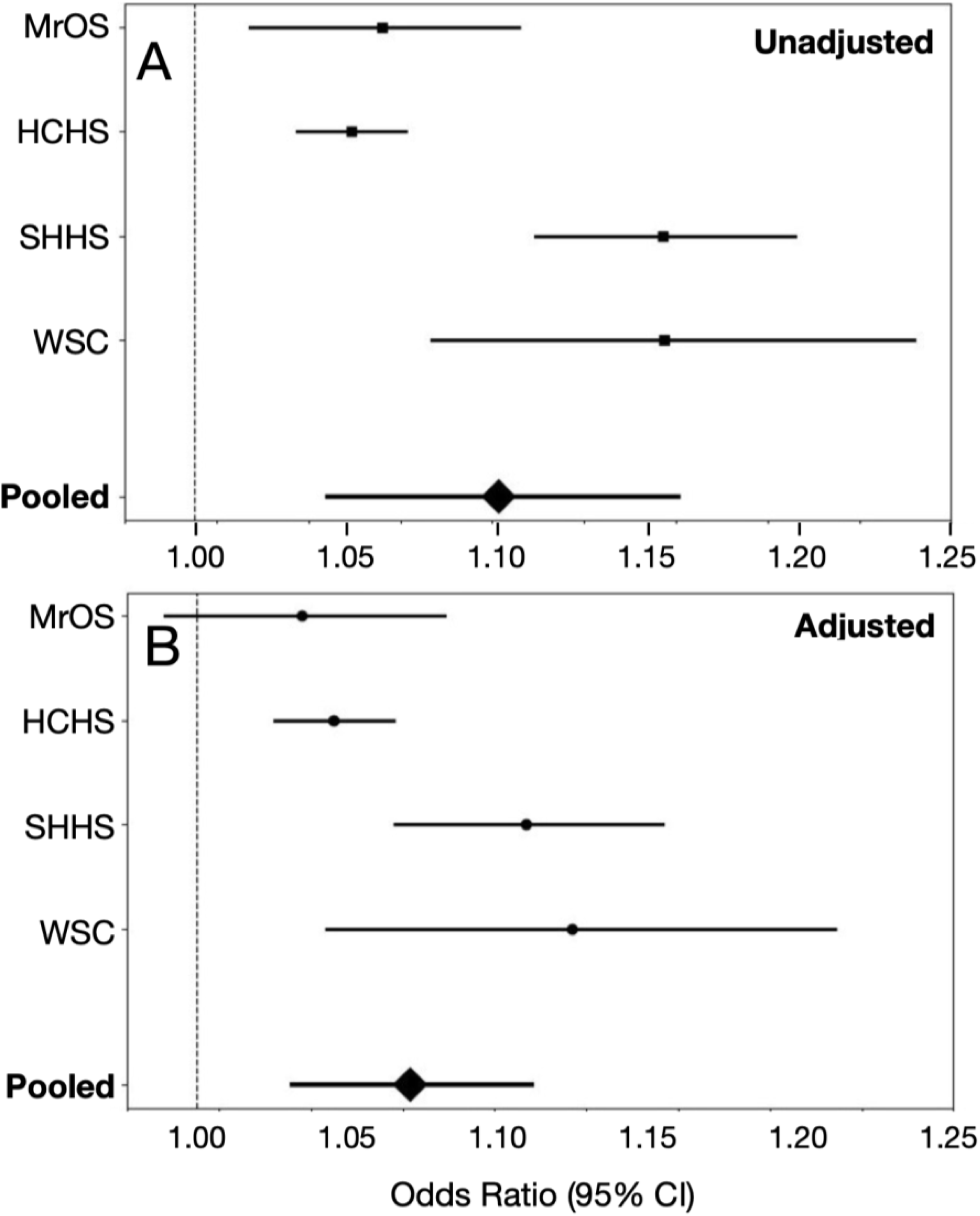
Forest plot illustrating the increase in odds of diabetes mellitus (DM) for each 1 brpm increase in nocturnal respiratory rate (NRR). **A)** Unadjusted (univariate) odds ratios (ORs) and 95% confidence intervals (CIs) for each dataset, along with the pooled estimate derived from random-effects meta-analysis. The pooled OR was 1.1 (95% CI: 1.04–1.16, P = 3.5 × 10⁻⁴). **B)** Adjusted (multivariate) ORs and 95% CIs for each dataset, along with the corresponding pooled estimate. The pooled OR was 1.07 (95% CI: 1.03–1.11, P = 3.0 × 10⁻⁴).

### Dichotomized NRR Analysis

In addition to evaluating NRR as a continuous predictor, we explored multiple dichotomization thresholds ranging from 15 to 21 brpm across all cohorts (see **Supplementary Fig. 1**). While odds ratios (ORs) and confidence intervals (CIs) varied by cohort and threshold, the univariate and multivariate associations between higher NRR and prevalent diabetes mellitus (DM) generally remained positive (OR > 1) across most cutoffs. For illustrative purposes, we report here the results for a representative cutoff of 18 breaths per minute (NRR ≥ 18 vs. < 18), selected arbitrarily from among the seven tested thresholds. In univariate models, this cutoff was significantly associated with increased odds of DM in SHHS (OR = 1.98, 95% CI: 1.55–2.51, *P* = 3.1 × 10⁻⁸), HCHS/SOL (OR = 1.36, 95% CI: 1.21–1.52, *P* = 1.2 × 10⁻⁷), and WSC (OR = 4.05, 95% CI: 2.48–6.63, *P* = 2.4 × 10⁻⁸), but not in MrOS (OR = 1.14, 95% CI: 0.86–1.50, *P* = 0.36).

After multivariable adjustment, the association remained statistically significant in SHHS (OR = 1.57, 95% CI: 1.22–2.03, *P* = 4.6 × 10⁻⁴), HCHS/SOL (OR = 1.31, 95% CI: 1.15–1.49, *P* = 3.8 × 10⁻⁵), and WSC (OR = 3.35, 95% CI: 1.93–5.83, *P* = 1.9 × 10⁻⁵), but not in MrOS (OR = 1.01, 95% CI: 0.75–1.34, *P* = 0.96). Although results were generated for all seven dichotomization thresholds, we report only those for the 18 breaths per minute cutoff here for brevity and consistency, rather than for any pre-specified biological or statistical reason.

### Sensitivity Analysis

Since a clear data-driven NRR dichotomization threshold was not apparent, we performed sensitivity analysis across NRR thresholds. **Figure 3** presents the results of a random-effects meta-analysis combining the dichotomized NRR analyses across cohorts. As the NRR threshold increases, the pooled odds ratio (OR) also tends to rise in both univariate and multivariate models, suggesting a dose–response relationship between elevated NRR and diabetes risk. Figure 3 presents the results of a random-effects meta-analysis pooling dichotomized NRR analyses across cohorts. As the NRR threshold increases, the corresponding pooled odds ratios (ORs) also rise in both univariate and multivariate models, suggesting a dose–response relationship between elevated NRR and diabetes risk. For a representative threshold of ≥18 brpm, the unadjusted pooled OR was 1.77 (95% CI: 1.23–2.55, *P* = 0.0022), while the adjusted pooled OR was 1.49 (95% CI: 1.10–2.02, *P* = 0.0098).

**Figure 3.**
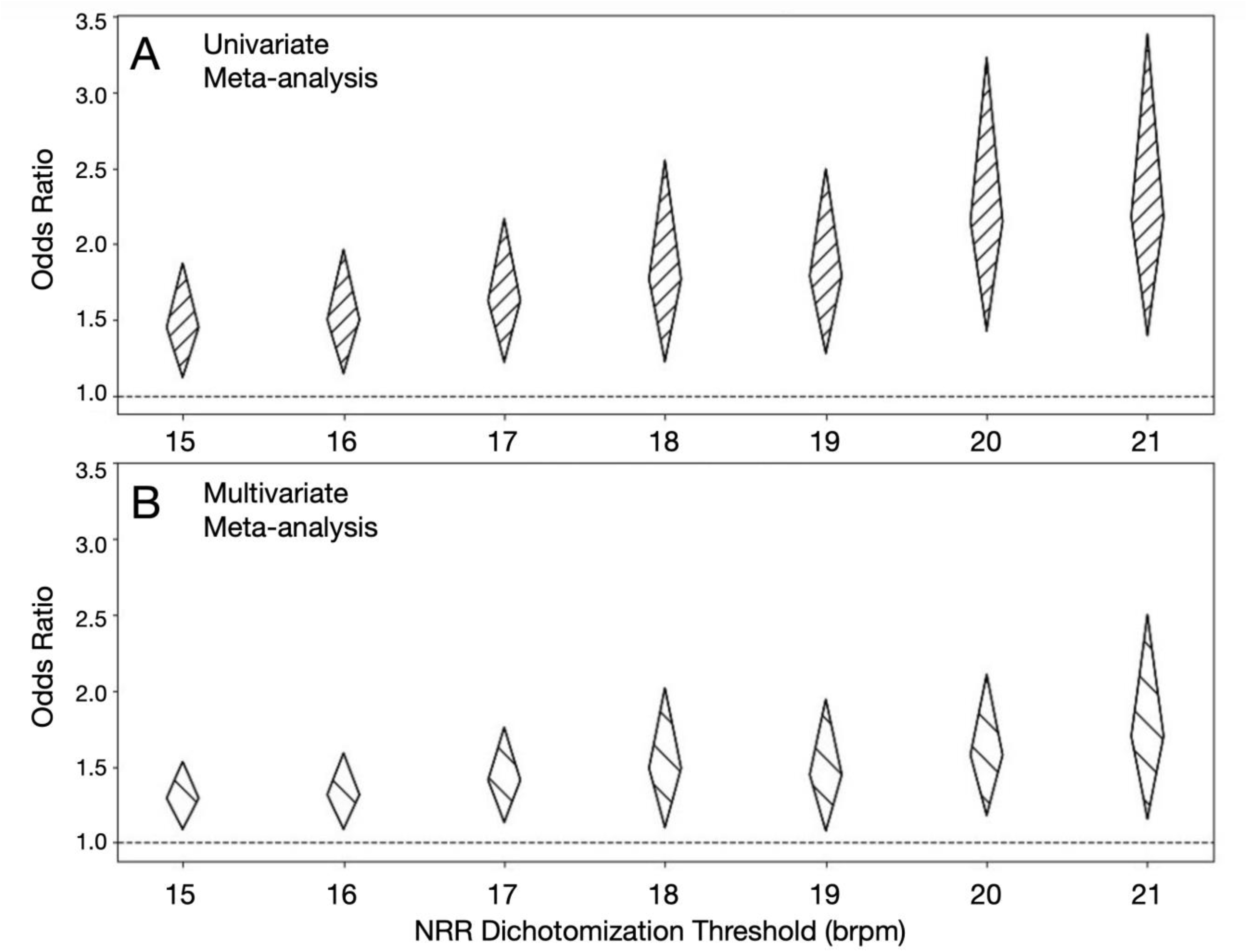
Sensitivity analysis for median NRR dichotomization threshold showing the resulting meta-analysis odds ratios at each threshold. The 18 brpm cutoff is shown here. All thresholds and their corresponding ORs and 95% CIs, are detailed in Supplemental Table 2. For the NRR = 18 brpm threshold, **A)** the unadjusted (univariate) pooled odds ratio was OR = 1.77 (95% CI 1.23–2.55), *P* = 0.0022, **B)** the adjusted (multivariate) pooled odds ratio was OR = 1.49 (95% CI 1.10–2.02), *P* = 0.0098.

## DISCUSSION

Respiratory rate is a fundamental vital sign that dynamically regulated by a central pattern generator within the brainstem based on inputs from chemoreceptors, the autonomic nervous system, voluntary effort and other higher order centers of the brain. Since DM causes autonomic neuropathy, we wondered whether respiratory rate might be associated with DM. If true, it may support respiratory monitoring as a convenient longitudinal biomarker of DM complications. Unfortunately, respiratory rate is not routinely measured outside of formal clinical settings. Therefore, we derived NRR from the raw waveforms of more than 25,000 elective overnight sleep studies from 4 large observational cohorts to investigate its relationship with DM in clinically stable outpatients. We found that for each breath-per-minute increase in median NRR, there was approximately a 10% increase in the odds of having diabetes mellitus (DM) in unadjusted models. This association remained statistically significant after adjusting for age, sex, BMI, smoking history, sleep apnea severity, and race, with an estimated 6% increase in odds per 1 brpm of NRR increase. Dichotomized analyses similarly showed higher odds of DM at elevated NRR groups, which was largely insensitive to choice of NRR threshold, from 15 to 21 brpm. At a representative threshold of 18 brpm, individuals had 77% higher odds of DM in unadjusted models and 49% higher odds of DM after multivariable adjustment.

Most studies of respiratory rate focus on acutely ill inpatients rather than clinically stable outpatients. In the hospital, respiratory rate is used to detect acute clinical decompensation and serves prominently in early warning scores. There, its relationship to DM focuses primarily on the rapid shallow breathing response to diabetic ketoacidosis. However, DM can affect respiratory physiology in more subtle ways in the outpatient setting. Unfortunately, the lack of routine outpatient monitoring leaves little high-quality respiratory data for analysis. Today, respiratory rate is only measured a few times per year at clinic visits by manually counting of breaths in a 15-30 second window in awake individuals. To overcome these barriers, we derived NRR from the chest belt waveforms collected during polysomnography sub-studies of large observational cohort studies and analyzed the relationship to DM. We had previously performed longitudinal respiratory monitoring using non-contact bed sensors and discovered that median NRR exhibits low night-to-night variability within individual patients even though variation of NRR between patients was high. This suggested that patient-level analysis of a single overnight sleep study might be sufficient to detect associations between NRR and determinants such as diabetes.

Although retrospective observational analyses cannot establish causality, there are multiple physiological pathways that may explain the observed association between DM and NRR. Individuals with DM often develop autonomic dysfunction and heightened sympathetic tone, which can increase respiratory rate. ^22-25^ In addition, DM has also been linked to impairments in pulmonary function and respiratory muscle strength, as demonstrated by reductions in forced expiratory volume in one second (FEV1), forced vital capacity (FVC), maximal inspiratory pressure (MIP), peak expiratory flow (PEF), diffusing capacity of the lungs for carbon monoxide (DLCO), and dynamic lung capacity.^25-29^ There are also DM-associated contributors to respiratory rate such as obesity and sleep-disordered breathing, however the DM-NRR associated persisted despite multivariate analysis adjusting for these factors.^30^ Given the independent and consistent association between NRR and DM, it is possible that NRR can serve as a dynamic biomarker for DM and its complications. Given the growing availability of non-contact respiratory monitoring devices during sleep, it will be increasing convenient to monitoring across time in an adherence-independent manner.

In this study, we densely sampled respiratory waveforms and quantified NRR at minute-scale epochs throughout the night. However, we only derived one median NRR for each night. While this summary measure was sufficient to establish the DM-NRR association, it overlooks valuable information about intranight variation and patterns that may include additional information. In addition, our study focused on breathing frequency but did not include information about relative amplitude or other modulations and patterns of breathing. Future work will focus on derivation and analysis of expanded feature sets to investigate respiratory dynamics across multiple time scales dynamically deeply phenotype DM-associated respiratory biomarkers in the outpatient setting.

Elevated mean NRR has been associated with an increased risk of all-cause and cardiovascular mortality in older adults.^31^ Additionally, nocturnal respiratory rate dynamics appear clinically relevant for predicting impending hospitalizations, assessing prognosis in heart failure, determining the benefit of implantable cardioverter-defibrillators (ICDs), and estimating the risk of non-sudden cardiac death in post-myocardial infarction patients.^32-37^ This further underscores the importance of longitudinal monitoring of respiratory physiology.

Strengths of our study include the long duration and high sampling density of our measurements as well as their collection during sleep, which maximizes data quality and avoids conscious and activity-dependent confounders. In addition, our findings derived from four well-characterized cohorts encompassing diverse populations, which improves generalizability. Moreover, using random-effects meta-analysis appropriately accounted for inter-study heterogeneity.

Our study also has several limitations. First, it is a retrospective analysis of observational cohorts. This can enable hypothesis-generating associations, but it cannot establish causality. Second, each patient PSG represented only a single night, often in a clinical lab, wired to many sensors, and as such, may not reflect the conditions and respiratory physiology of a typical night of sleep at home. Third, although multiple covariates were adjusted for, residual confounding remains possible, particularly given slight differences in the definitions of available covariates across cohorts. Finally, since there is no established NRR threshold for dichotomization analysis, we were left to perform sensitivity analysis across a range of cutoffs. Nevertheless, the consistency of our findings across diverse and large cohorts provides important support for the relationship between DM and NRR.

Future research should examine the mechanism underlying the DM-NRR association as well as its association with DM severity, management, reversibility, and hard outcomes. If NRR proves to be a high-quality dynamic biomarker of DM-complications, it is attractive for longitudinal monitoring via the growing number of non-contact and adherence-independent bed sensors.

## Data Availability

All data produced in the present work are contained in the manuscript

## ACKNOWLEDGEMENTS

The work was funded by the American Heart Association (AHA) Predoctoral Fellowship 26PRE1560767 (R.P.V.). NIH T32 Training Program on Cardiovascular Physiology and Pharmacology Program (D.T.B.); NIH NHLBI R33 HL168785 (K.R.K.), DOD HT94252410105 (K.R.K.), Wellcome Leap In Utero Program. The Sleep Heart Health Study (SHHS) was supported by National Heart, Lung, and Blood Institute cooperative agreements U01HL53916 (University of California, Davis), U01HL53931 (New York University), U01HL53934 (University of Minnesota), U01HL53937 and U01HL64360 (Johns Hopkins University), U01HL53938 (University of Arizona), U01HL53940 (University of Washington), U01HL53941 (Boston University), and U01HL63463 (Case Western Reserve University). The National Sleep Research Resource was supported by the National Heart, Lung, and Blood Institute (R24 HL114473, 75N92019R002). The Hispanic Community Health Study/Study of Latinos (HCHS/SOL) was performed as a collaborative study supported by contracts from the NHLBI to the University of North Carolina (N01-HC65233), University of Miami (N01-HC65234), Albert Einstein College of Medicine (N01-HC65235), Northwestern University (N01-HC65236), and San Diego State University (N01-HC65237) (AG05407, AR35582, AG05394, AR35584, AR35583, AG08415). The National Sleep Research Resource was supported by the National Heart, Lung, and Blood Institute (R24 HL114473, 75N92019R002). The National Heart, Lung, and Blood Institute provided funding for the ancillary MrOS Sleep Study, "Outcomes of Sleep Disorders in Older Men," under the following grant numbers: R01 HL071194, R01 HL070848, R01 HL070847, R01 HL070842, R01 HL070841, R01 HL070837, R01 HL070838, and R01 HL070839. The National Sleep Research Resource was supported by the National Heart, Lung, and Blood Institute (R24 HL114473, 75N92019R002). This Wisconsin Sleep Cohort Study was supported by the U.S. National Institutes of Health, National Heart, Lung, and Blood Institute (R01HL62252), National Institute on Aging (R01AG036838, R01AG058680), and the National Center for Research Resources (1UL1RR025011). The National Sleep Research Resource was supported by the U.S. National Institutes of Health, National Heart Lung and Blood Institute (R24 HL114473, 75N92019R002).

## HUMAN SUBJECTS

The retrospective analyses of observational data were considered exempt per Institutional Review Board protocol #810998.

## INTERESTS

Dr. King and Nicholas Harrington are inventors on a patent application describing the Bedscales technology. Dr. King is Founder, Director, on Board of Directors and holds equity in Nightingale Labs. Dr. Harrington is an employee of Nightingale Labs.

---

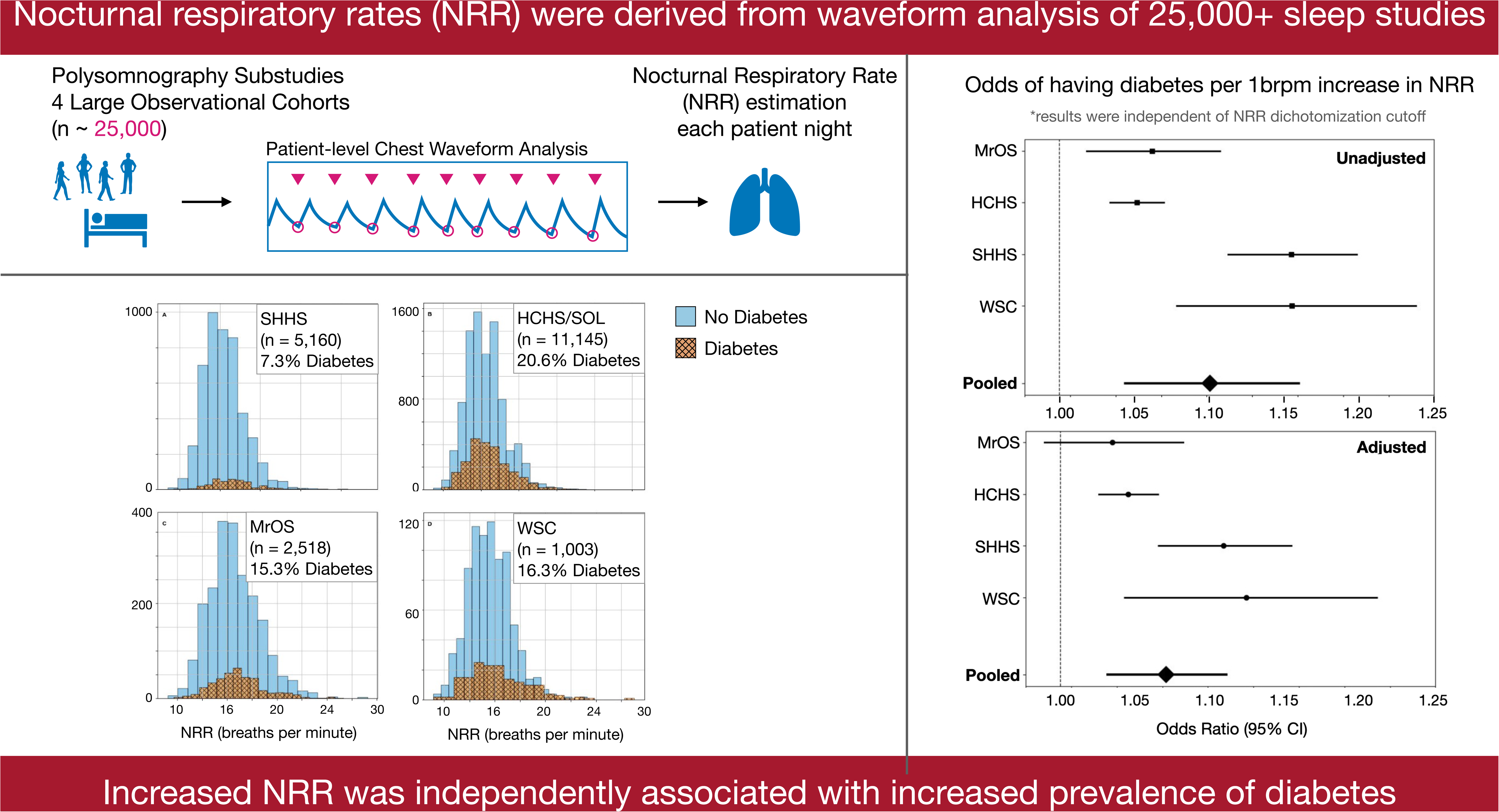

