## Supplemental Information for "Diabetes is associated with increased nocturnal respiratory rate"

**Data and Resource Availability**

The data used in this study is available from the National Sleep Research Resource (NSRR) upon request. <https://sleepdata.org/>

| **Cutoff**  **(brpm)** | **Pooled**  **OR** | **95% CI Lower** | **95% CI Upper** | **P-value** | **Analysis Type** |
| --- | --- | --- | --- | --- | --- |
| 15 | 1.453 | 1.125 | 1.876 | 0.0042 | Univariate |
| 16 | 1.504 | 1.151 | 1.965 | 0.0028 | Univariate |
| 17 | 1.629 | 1.224 | 2.169 | 0.00082 | Univariate |
| 18 | 1.77 | 1.228 | 2.552 | 0.0022 | Univariate |
| 19 | 1.791 | 1.284 | 2.497 | 0.00059 | Univariate |
| 20 | 2.15 | 1.431 | 3.231 | 0.00023 | Univariate |
| 21 | 2.179 | 1.403 | 3.385 | 0.00053 | Univariate |
| 15 | 1.293 | 1.09 | 1.534 | 0.00313 | Multivariate |
| 16 | 1.316 | 1.09 | 1.59 | 0.00432 | Multivariate |
| 17 | 1.412 | 1.133 | 1.761 | 0.00213 | Multivariate |
| 18 | 1.491 | 1.101 | 2.019 | 0.00977 | Multivariate |
| 19 | 1.449 | 1.08 | 1.944 | 0.01346 | Multivariate |
| 20 | 1.577 | 1.18 | 2.109 | 0.00210 | Multivariate |
| 21 | 1.702 | 1.157 | 2.502 | 0.00687 | Multivariate |

**Supplementary Table 1: The odds of DM for high NRR vs. low NRR groups**, dichotomized at variable thresholds. The results indicate that the high NRR group has significantly higher odds of DM irrespective of dichotomization thresholds. In the main manuscript, the ORs at 18brpm are reported.
